# Coagulation abnormalities in children with uncorrected congenital heart defects seen at a teaching hospital in a developing country

**DOI:** 10.1101/2022.01.31.22270219

**Authors:** Omotola O. Majiyagbe, Adeseye M. Akinsete, Titilayo A. Adeyemo, Ekanem N. Ekure, Christy A. N. Okoromah

**Author notes:** Corresponding author (OM). Department of Paediatrics, Massey Street Children’s hospital, Lagos Island, Lagos, Nigeria. **Contributor Role**. **Omotola O. Majiyagbe** ROLES: Conceptualization, Data curation, Formal analysis, Investigation, Methodology, Project administration, Resources, Writing - original draft, Writing – review & editing. **Adeseye M. Akinsete** ROLES: Methodology, Project administration, Supervision, Writing – review & editing. **Titilayo A. Adeyemo** ROLES: Methodology, Project administration, Resources, Supervision, Validation, Writing – review & editing. **Ekanem N. Ekure** ROLES: Conceptualization, Methodology, Project administration, Resources, Supervision, Writing – original draft, Writing – review & editing. **Christy A. N. Okoromah** ROLES: Supervision, Writing-review & editing. Competing interests: The authors have declared that no competing interests exist.

## Abstract

**Background:** Coagulation abnormality is a significant complication and cause of mortality in children with uncorrected congenital heart defects (CHD). The aim of this study was to determine the prevalence of coagulation abnormalities and the associated factors in children with uncorrected CHD.

**Method:** A cross sectional study conducted to determine the prevalence of coagulation abnormalities among 70 children with uncorrected CHD aged six months to 17 years and 70 age and sex matched apparently healthy controls. Coagulation abnormality was determined using complete blood count, prothrombin time, activated partial thromboplastin time and D-dimer assay.

**Results:** The prevalence of coagulation abnormalities among children with CHD and controls was 37.1% and 7.1% respectively. Children with Cyanotic CHD had a significantly higher prevalence of coagulation abnormalities compared to children with Acyanotic CHD (57.1% versus 17.1%). Haematocrit and oxygen saturation levels were significantly associated with coagulation abnormalities.

**Conclusion:** This study affirms that coagulation abnormality is high in children with uncorrected CHD. Oxygen saturation and haematocrit are factors of coagulation abnormalities. Routine coagulation screening is recommended especially in children with cyanotic congenital heart defects to improve their quality of life and reduce morbidity and mortality while awaiting definitive surgeries.

## Introduction

Globally, congenital heart defect (CHD) defined as a structural and functional defect of the heart present at birth, affecting the heart or adjacent great blood vessels, detected either at birth or later in life, accounts for nearly a third of all major congenital anomalies with an estimated prevalence of 8-12 per 1000 live births.[1,2,3]

In Nigeria, the estimated prevalence varies from 4.6 - 9.3 per 1000 children.[4,5] The prevalence of uncorrected CHD, growing to adulthood in Nigeria is unknown, however, in some developed countries like United States, 85% of children with CHD are documented to survive into adulthood since the advent of neonatal repair of complex heart lesions.[6] A multi-center hospital survey in Nigeria by Ekure *et al* [7] in 2017 reported that 84% of the enrolled 1,296 children with CHD still had uncorrected lesions. The delay in corrective surgery was due to late presentation, inadequate infrastructure as well as a high cost of cardiac care. Morbidity from uncorrected CHD may be due to anaemia, infections, malnutrition, congestive cardiac failure, coagulation abnormalities, cerebrovascular accident, bleeding diathesis and pulmonary hypertension, which can have a devastating effect on their quality of life.[8]

Coagulation defined as a physiologic process of maintaining haemostasis in the body, can be disrupted in children with CHD leading to functional and structural abnormalities. This is usually due to the chronic state of hypoxia in CCHD as well as chronic heart failure in children with ACHD. These mechanisms lead to compensatory erythrocytosis, hyperviscosity, thrombocytopaenia, platelet aggregation suppression, factor deficiencies, fibrinolysis and thromboembolic phenomenon.[9] Most cases of coagulation abnormalities are mild and asymptomatic, however, severe cases can lead to major complications such as cerebrovascular accidents, limb loss from ischaemia and disseminated intravascular coagulopathy (DIC) which may worsen morbidity; the effects of which may persist even after correction of the heart defects.[10]

Coagulation abnormalities can be assessed using the baseline tests for detecting coagulation abnormalities [Prothrombin time (PT), activated partial thromboplastin time (APTT)] in addition to D-dimer assay which has a 95% sensitivity for detecting disseminated intravascular coagulation (DIC).[11]

Data is limited concerning the prevalence and significance of CHD coagulation abnormality in developing countries. Studies from developed countries reported a wide prevalence of 19 to 70% [12,13], thus extrapolating these findings to populations in developing countries might be misleading. This is because the spectrum of routine care and affordability of interventions readily available in developed countries is mostly unavailable in developing countries like Nigeria. Furthermore, coagulopathy in CCHD has been studied more frequently than ACHD, although acyanotic CHD is commonly seen. [14] Therefore, to improve the quality of care offered to this population until definitive corrective intervention is obtained, it is important to identify, monitor and treat any coagulation abnormality to avoid development of irreversible life-limiting end-organ damage.

Thus, the objective of the study was to determine the prevalence of coagulation abnormality in the paediatric uncorrected congenital heart defect population and to identify the associated risk factors in a cohort of Nigerian children.

## Materials and methods

It was a cross sectional study conducted at the Lagos University Teaching Hospital (LUTH), Lagos, South west Nigeria from March 2019 to September 2019, after ethical approval was obtained from the Health Research Ethics Committee. The institution is a tertiary hospital that receives referrals from other healthcare facilities in the state and its environs.

Study participants were recruited from the Cardiology clinic, Cardiovascular laboratory and Paediatric outpatient clinics of the hospital using a consecutive sampling method.

### Study population

The presence of congenital heart defect was based on the documentary evidence of echocardiography report. Thereafter, consecutive patients with uncorrected CHDs between 6 months to 17 years of age were included in the study while healthy age and sex matched children, who had no clinical evidence of congenital heart defects were recruited as controls. Written informed consent was obtained from parents or guardians of all study participants with additional informed written assent from children aged seven years and above.

Patients previously diagnosed with chronic renal failure, chronic liver disease, malignancies, HIV infection, sickle cell anaemia were excluded from the study. Children who had been on anti-inflammatory medications such as aspirin in the preceding two weeks, as well those with family history of bleeding disorders such as haemophilia were also excluded. Clinical examination and anthropometry were carried out on CHD subjects and controls and a pre-tested questionnaire (S1 Appendix) was also used to obtain biodata, clinical information, and socio-demographic characteristics (age, gender, socioeconomic class which included occupational status and highest level of education attained by each parent and/or guardian using Oyedeji classification of social class). [15] Heart failure was classified using the Modified Ross Heart Failure Classification for Children.

Prothrombin time (PT) and APTT values greater than 2SD of the mean values were reported as prolonged, as recommended by the Clinical & Laboratory Standard Institute (CLSI) on defining, establishing and verifying reference intervals in the clinical laboratory.[16]

### Blood sample collection

Four milliliters of venous blood was withdrawn aseptically from each participant; two milliliters was added into a marked 3.2% citrated bottle for the clotting profile analysis (so as to maintain a standard 9:1 blood to anticoagulant ratio), while two milliliters was collected into an ethylene diamine tetra-acetic acid (EDTA) bottle for evaluation of haematocrit and platelet count. For cyanotic patients with known haematocrit levels above 55%, the haematocrit value was first checked and the amount of anticoagulant in the collection tube was adjusted according to the CLSI guidelines.[16]

All blood samples withdrawn were transported in a cooler box (+4 to +8 °C) in an upright position on the tube racks to the Central Research Laboratory of the College of Medicine, University of Lagos (CMUL), Lagos. Before receiving the samples at the LUTH Central Research Laboratory, the haematologist independently inspected the collected samples, and removed those samples that were deemed insufficient, with blood clots or evidence of haemolysis. Within 4 hours of collection, the samples were centrifuged at 1500 rpm for 15mins, the resultant platelet poor plasma was extracted using pipettes into cryo bottles in three aliquots of 0.5mls and stored at -70 °C at the CMUL Central Research Laboratory for a month to maintain potency until analysis.

### Laboratory analysis

Complete blood count was analysed using a Mindray BC-3200 Automated Haematology Analyzer (Shenzhen, China).

Activated partial thromboplastin time was determined using the coagulometer, a CA-101 (Sysmex, India), a compact semi-automated coagulation analyzer. Fifty microliters of platelet poor plasma (0.05mls) was pipetted into a cuvette and fifty microliters of APTT reagent, Actin FS (Siemens Healthcare Diagnostics Product GmbH, Marburg, Germany) containing phospholipid and a contact activator, was added and incubated in the coagulometer at 37 °C for three minutes. Thereafter, 0.05mls of calcium chloride solution (pre-warmed to 37 °C) was added and timing begun. The aPTT is the time taken from the addition of the calcium to the formation of a fibrin clot.

Prothrombin time was also determined using the coagulometer. Fifty microliters of platelet poor plasma (0.05mls) was pipetted into a cuvette and incubated for 30 seconds. Thereafter, 0.1ml of PT reagent, Dade Innovin (Siemens Healthcare Diagnostics Product GmbH, Marburg, Germany) a tissue factor containing phospholipid, was added and incubated at 37°C and timing begun.

All tests were done in duplicates, and the average of the two readings was recorded, a difference of not more than 10% was allowed. A known quality control sample (pooled plasma of known PT and APTT values) was also run to ascertain validity of the test.

D-dimer was assayed by enzyme linked immunosorbent assay (ELISA) using the Human D-dimer ELISA microwells kit (Bioassay Technology Laboratory, China) designed for quantitative detection of d-dimer in human serum or plasma. The kit is a solid phase sandwich assay method, based on a streptavidin-biotin principle. Coagulation abnormality was defined as thrombocytopaenia, prolonged prothrombin time and/or activated partial thromboplastin time and elevated D-dimer assay levels.

### Follow up of study participants

Laboratory results were made available to study participants. This was done mainly by telephone calls and distributing personal hard copies of the laboratory results to the parents/guardian during their follow up appointments. For the children with abnormal but asymptomatic coagulation tests, results were communicated to their managing physicians, to be co-managed with the Haematology unit of LUTH through short clinic appointments with a recommendation for proactive vigilance for a bleeding diathesis or thrombotic event or both. The caregivers of children with uncorrected CHD were also counseled on the need for prompt surgical correction of the cardiac defects.

### Data analysis

Statistical analysis was performed using Statistical Package for Social Sciences (SPSS) (version 21) Armonk, NY:IBM Corp.

Quantitative data was tested for normality using Shapiro-Wilk test. Normally distributed data was summarized using mean and standard deviation, while skewed data was presented as median and interquartile range. Conversely, categorical variables (demographic, anthropometry and laboratory findings) were summarized using frequencies and proportions. Comparison of the prevalence of coagulation abnormalities between the subject group and control group was done using the Chi square test. Comparison of normally distributed continuous data (laboratory findings) between the cyanotic and acyanotic groups was tested using student t test while similar comparison between skewed data were made using Mann Whitney U test. Comparison of categorical variables between cyanotic and acyanotic groups was done using the Chi-square test. Variables that were statistically significantly associated with coagulation abnormalities were subjected to further statistical test using multiple logistic regression to determine the independent predictors of coagulation abnormalities. For all statistical analyses, p value less than 0.05 was considered statistically significant.

## Results

A total of 144 children were recruited for the study. Four of them were excluded from data analysis on account of inability to run the laboratory tests due to clotted samples.

### Characteristics of CHD subjects and the controls

A total of one hundred and forty children were enrolled in the study: seventy children each in the uncorrected congenital heart defects and apparently healthy aged and sex matched control groups.

Table 1 shows a summary of the demography and anthropometric findings in CHD subjects and controls. The study population ages ranged from 6 months to 15 years with majority in the 1-5 years group. The median (IQR) age of the two groups was comparable 36.0 (18.0-84.0) months in the CHD and 48.0 (20.0-87.0) months in the healthy control (p-value = 0.985). The male-female proportion was 42.9% and 57.1%. Most of the caregivers of the study population were in the middle socioeconomic class. The median weight, height and body mass index were lower for CHD subjects compared to controls. Children with CHD were significantly stunted and wasted compared to the healthy controls. Twenty-nine (41.4%) and 26 (37.2%) of the children with uncorrected CHDs were wasted and stunted respectively. compared to 11 (15.7%) and 10 (14.3%) of the healthy controls who were wasted and stunted, p=<0.001

**Table I:** Socio-demographic and anthropometry of study population

| Variable<br>N (%) | Congenital heart defects | Controls | Test<br>statistic | $p$-value |
| --- | --- | --- | --- | --- |
| | n=70 | n=70 | $\chi^2$ | |
| Age, median (IQR), months | 36.0(18.0-84.0) | 48.0(20.0-87.0) |  | 0.985 |
| **Age group** |  |  |  |  |
| 6 months – 1 year | 8(11.4) | 8(11.4) |  |  |
| 1 to <5 years | 34(48.6) | 34(48.6) | $\chi^2=0.000$ | 1.000 |
| 5 to <10 years | 19(27.1) | 19(27.1) |  |  |
| $\geq 10$ years | 9(12.9) | 9(12.9) | | |
| **Sex** |  |  |  |  |
| Females | 40(57.1) | 40(57.1) | $\chi^2=0.116$ | 1.000 |
| Males | 30(42.9) | 30(42.9) |  |  |
| **Socioeconomic class** |  |  |  |  |
| Low | 11 (15.7) | 4 (5.7) |  |  |
| Middle | 35 (50.0) | 29 (41.4) | $\chi^2=6.600$ | 0.037 |
| High | 24 (34.3) | 37 (52.9) |  |  |
| **Anthropometry** |  |  |  |  |
| BMI for age, median (IQR) | -1.6 (-2.7 to -0.7) | -0.4(-1.0 to -0.9) | F=35.169 | <0.001<sup>#</sup> |
| Height for age, median (IQR) | -1.4 (-2.4 to -0.5) | 0 (-1.3 to 0.8) | F=24.669 | <0.001<sup>#</sup> |
| Weight for age, median (IQR) | -2.0 (-3.0 to -1.3) | -0.4 (-0.9 to 0.4) | F=60.682 | <0.001<sup>#</sup> |
| **BMI for age, n (%)** |  |  |  |  |
| Normal | 41 (58.6) | 52 (74.3) | F=23.612 | <0.001 |
| Wasted | 13 (18.5) | 10 (14.3) |  |  |
| Severely wasted | 16 (22.9) | 1 (1.4) |  |  |
| Overweight | 0 (0) | 7 (10) |  |  |
| **Height for age, n (%)** |  |  |  |  |
| Normal | 44 (62.8) | 55 (78.6) | F=17.845 | <0.001 |
| Stunted | 13 (18.6) | 9 (12.9) |  |  |
| Severely stunted | 13 (18.6) | 1 (1.4) |  |  |
| Tall | 0 (0) | 5 (7.1) |  |  |

$\chi^2$ : Chi-square. F= Fischer's test
<sup>#</sup>p value statistically significant

### Characteristics of children with CHD

For children with acyanotic congenital heart defects: Isolated VSD 14 (40%), Isolated ASD 6 (17.1%) and Isolated PDA 3 (8.5%) were the most common types, while Tetralogy of Fallot 20 (57.1%) and Truncus arteriosus 4 (11.4%) were the commonest CCHD. Heart failure was diagnosed in 32 of the 70 CHD children, of which 87.5% of them had ACHD (Modified Ross Heart failure classification) as shown in Table 2.

**Table 2:**
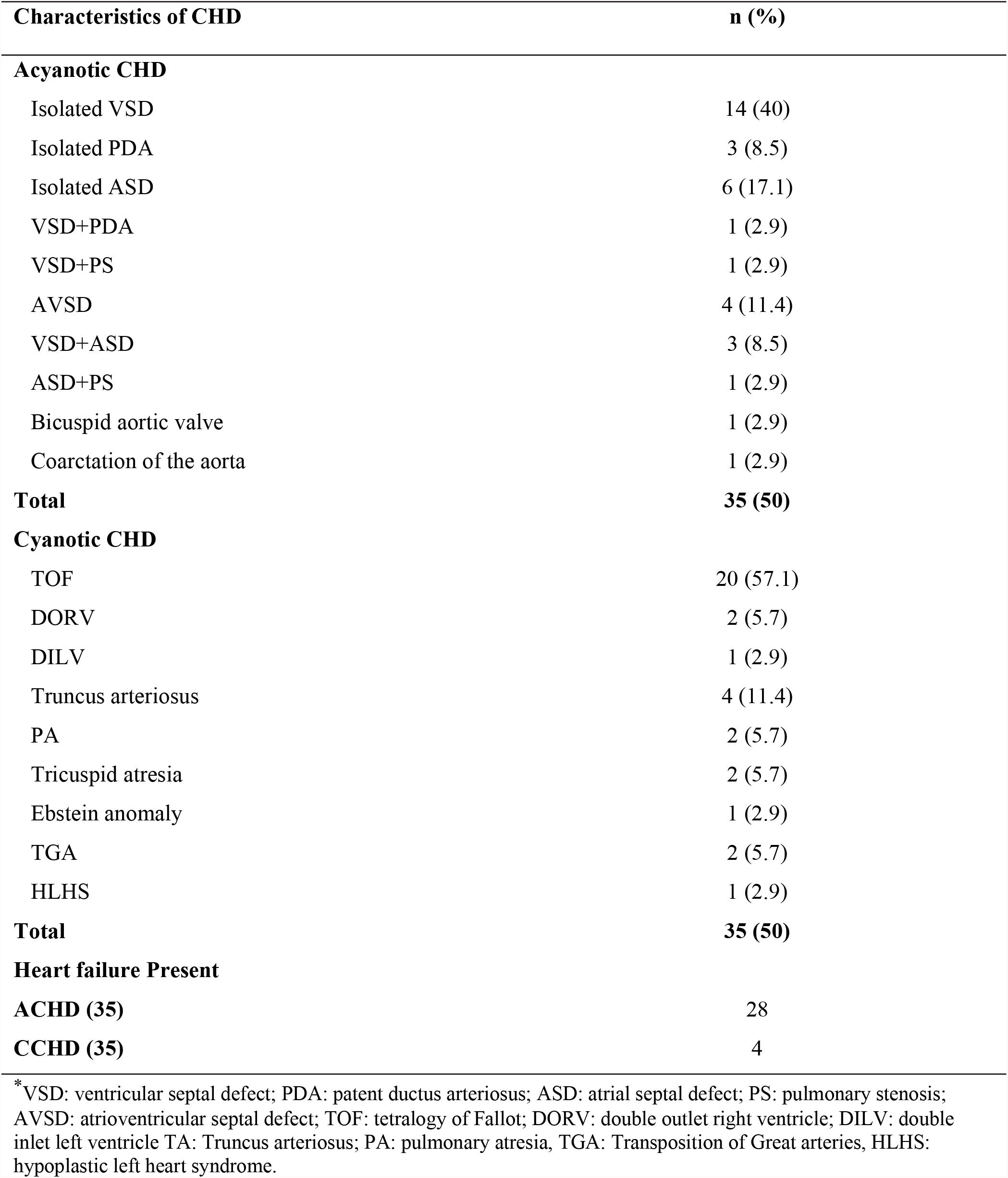
Distribution of Congenital heart defects.

| Characteristics of CHD | n (%) |
| --- | --- |
| **Acyanotic CHD** |  |
| Isolated VSD | 14 (40) |
| Isolated PDA | 3 (8.5) |
| Isolated ASD | 6 (17.1) |
| VSD+PDA | 1 (2.9) |
| VSD+PS | 1 (2.9) |
| AVSD | 4 (11.4) |
| VSD+ASD | 3 (8.5) |
| ASD+PS | 1 (2.9) |
| Bicuspid aortic valve | 1 (2.9) |
| Coarctation of the aorta | 1 (2.9) |
| **Total** | **35 (50)** |
| **Cyanotic CHD** |  |
| TOF | 20 (57.1) |
| DORV | 2 (5.7) |
| DILV | 1 (2.9) |
| Truncus arteriosus | 4 (11.4) |
| PA | 2 (5.7) |
| Tricuspid atresia | 2 (5.7) |
| Ebstein anomaly | 1 (2.9) |
| TGA | 2 (5.7) |
| HLHS | 1 (2.9) |
| **Total** | **35 (50)** |
| **Heart failure Present** |  |
| **ACHD (35)** | 28 |
| **CCHD (35)** | 4 |

\*VSD: ventricular septal defect; PDA: patent ductus arteriosus; ASD: atrial septal defect; PS: pulmonary stenosis; AVSD: atrioventricular septal defect; TOF: tetralogy of Fallot; DORV: double outlet right ventricle; DILV: double inlet left ventricle TA: Truncus arteriosus; PA: pulmonary atresia, TGA: Transposition of Great arteries, HLHS: hypoplastic left heart syndrome.

### Prevalence of Coagulation abnormalities in children with CHD and Controls

The proportion of uncorrected CHD cohort with coagulation abnormality, defined as thrombocytopaenia, prolonged prothrombin time and/or activated partial thromboplastin time and elevated D-dimer assay was 26 (37.1%), which was much higher than 5 (7.1%) in apparently healthy controls as shown in Fig 1.

**Fig 1.**
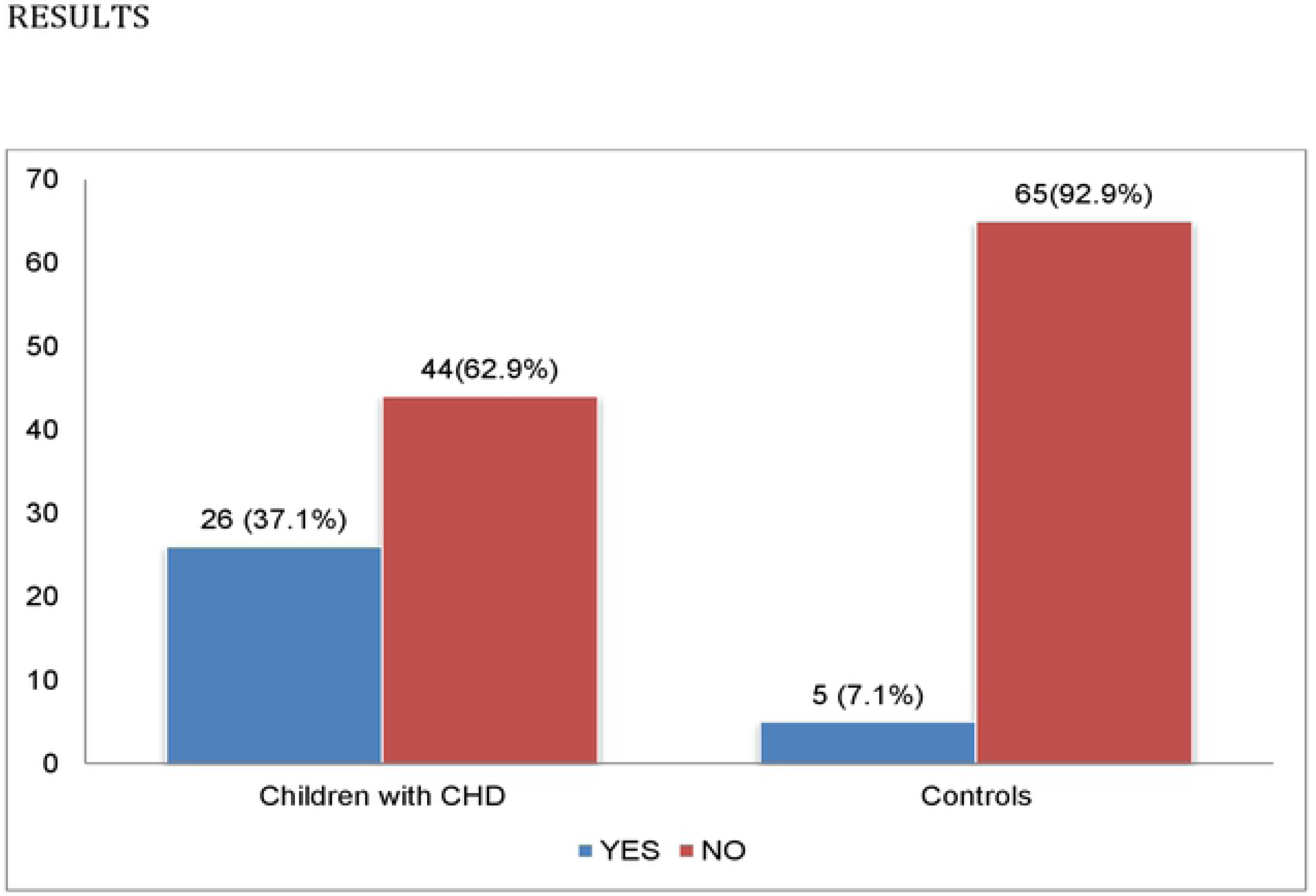
Prevalence of coagulation abnormalities in children with uncorrected CHD and controls

### Comparison of Coagulation profile in children with CHD and Controls

Table 3 shows the mean values of coagulation parameters in CHD subjects and controls. Children with CHD had prolonged APTT and PT (44.4 *±* 10.7 and 14.7 *±* 3.5 respectively) compared with controls (39.7 *±* 7.3 and 12.9 ± 1.7) (p-value <0.05). The median platelet count of the children with congenital heart defects was much lower than the median of the controls. However, there was no significant difference in the D-dimer levels between children with CHD and controls.

**Table 3:** Comparison of coagulation profile between CHD subjects and controls

| Variables | Congenital heart defects | Controls | Test statistics | P value |
| --- | --- | --- | --- | --- |
| **Median (IQR) platelet count**<br>($\times 10^9$ cells/mm<sup>3</sup>) | 240.1 (163.0-326.0) | 356.0(275.0-679.0) | U | 0.000 |
| **Prothrombin time,**<br>*mean ± SD, seconds* | 14.7±3.5 | 12.9±1.7 | t | <0.001 |
| **International Normalized Ratio**<br>*mean ± SD* | 1.1±0.3 | 0.9±0.1 | t | <0.001 |
| **Activated partial thromboplastin time,**<br>*mean ± SD, seconds* | 44.4±10.7 | 39.7±7.3 | t | 0.003 |
| **Median (IQR) D-dimer (pg/ml)** | 1.0(0.8-1.5) | 1.2(0.8-2.1) | U | 0.092 |

U: Mann Whitney U test t: Student's t-test

### Prevalence of Coagulation abnormalities in children with Acyanotic and Cyanotic Congenital heart defects

Coagulation abnormalities were more common in the cyanotic CHD group compared to the acyanotic group [20 (57.1%) versus 6 (17.1%)] with a p value of 0.001. The proportion of coagulation abnormality in the two subgroups is as shown in Table 4 Six children with ACHD had coagulation abnormalities: 5 had abnormal results in only one test while 1 had in two tests.

**Table 4:** Coagulation abnormalities in children with ACHD and CCHD

| **Variable**<br>**N (%)** | **ACHD**<br>**n=35** | **CCHD**<br>**n=35** | **Test statistic**<br>**$\chi^2$** | **p-value** |
| --- | --- | --- | --- | --- |
| Coagulation abnormality | 6 (17.1) | 20 (57.1) | 11.933 | 0.001 |
| Thrombocytopenia | 0(0.0) | 8(22.9) | 9.032 | 0.002 |
| Prolonged PT/INR | 2(5.7) | 11(31.4) | 7.652 | 0.006 |
| Prolonged APTT | 2(5.7) | 9(25.7) | 5.285 | 0.023 |
| Elevated D-dimer (pg/ml) | 3(8.6) | 3(8.6) | 0.000 | 0.663 |

$\chi^2$ : Chi-square.

Twenty children with CCHD had coagulation abnormalities: 12 had abnormal results in one test while 8 had in two or more tests.

### Factors associated with coagulation abnormalities in children with CHD

There was no significant association between the socio-demographic characteristics (age, sex, and socioeconomic class), nutritional status (height for age, BMI) and the prevalence of coagulation abnormality in children with congenital heart defects. Coagulation abnormality was more in CHD subjects that were less than 5 years of age, though it was not statistically significant. CHD subjects with coagulation abnormality had higher mean values of haematocrit and lower mean values of oxygen saturation levels as shown in Table 5.

**Table 5:** Association between socio demographic characteristics, nutritional status and the prevalence of coagulation abnormality in children with CHD.

| Variables (n=70) | Coagulation abnormality |  | Test statistics | P-value |
| --- | --- | --- | --- | --- |
|  | *Yes (n=26)* | *No (n=44)* |  |  |
| **Age group** |  |  |  |  |
| 6 months to <1 year | 3(11.5) | 5(11.4) |  |  |
| 1 year to <5 years | 13(50.0) | 21(47.7) | $\chi^2=1.076$ | 0.783 |
| 5 years to <10years | 8(30.8) | 11(25.0) |  |  |
| $\geq 10$ years | 2(7.7) | 7(15.9) | | |
| **Sex** |  |  |  |  |
| Male | 11(42.3) | 19(43.2) | $\chi^2=0.193$ | 0.660 |
| Female | 15(57.7) | 25(56.8) |  |  |
| **Height for age** |  |  |  |  |
| Normal | 16(61.6) | 28(63.6) |  | 0.985 |
| Stunted | 5(19.2) | 8(18.2) | $\chi^2=0.031$ | |
| Severely stunted | 5(19.2) | 8(18.2) |  |  |
| **BMI for age** |  |  |  |  |
| Normal | 13(50.0) | 28(63.6) | $\chi^2=1.661$ | |
| Wasted | 5(19.2) | 8(18.2) |  | 0.436 |
| **Heart failure class** |  |  |  |  |
| None | 13(50.0) | 25(56.8) |  |  |
| Class I | 8(30.8) | 18(40.9) |  | 0.051 |
| Class II | 5(19.2) | 0(0.0) |  |  |
| Class III | 0(0.0) | 1(2.3) |  |  |
| **Oxygen saturation in room air (mean ± SD)** | 79.6 ± 11.3 | 89.8 ± 10.6 | t=14.202 | 0.000 |
| **Haematocrit (mean ± SD)** | 46.5 ± 14.5 | 38.8 ± 10.7 | t=6.541 | 0.014 |

$\chi^2$ : Chi-square. t=Student t-test

Table 6 showed that socio-demographic characteristics (age, sex) and anthropometry were not independent predictors of coagulation abnormalities. Coagulation abnormality was 6 times [6.444 (2.135-19.456)] more likely to occur in children with CCHD compared with children with ACHD.

**Table 6:** Independent variables associated with coagulation abnormality

| Variable | Adjusted odds ratio | P value | 95% CI |  |
| --- | --- | --- | --- | --- |
|  |  |  | Lower bound | Upper bound |
|  |  |  | **Coagulation profile** |  |
| **Age group** |  |  |  |  |
| 6 months – 1 year (vs. $\geq 10$ years) | 2.100 | 0.494 | 0.251 | 17.594 |
| 1 to <5 years (vs. $\geq 10$ years) | 2.167 | 0.377 | 0.389 | 12.063 |
| 5 to <10 years (vs. $\geq 10$ years) | 2.545 | 0.313 | 0.414 | 15.652 |
| **Sex** |  |  |  |  |
| Male vs. Female | 1.254 | 0.660 | 0.469 | 3.309 |
| **Cardiac defects** |  |  |  |  |
| Cyanotic CHD vs Acyanotic | 6.444 | 0.001 | 2.135 | 19.456 |
| **Height for age** |  |  |  |  |
| Normal (vs. severely stunted) | 1.000 | 1.0000 | 0.206 | 4.856 |
| Stunted (vs. severely stunted) | 0.914 | 0.890 | 0.255 | 3.272 |
| **BMI for age** |  |  |  |  |
| Normal (vs. severely wasted) | 0.464 | 0.203 | 0.143 | 1.511 |
| Wasted (vs. severely wasted) | 0.625 | 0.535 | 0.141 | 2.763 |

Table 7 showed that haematocrit level and oxygen saturation had a statistically significant association on levels of platelet count, prothrombin time and INR. The model shows that haematocrit value and oxygen saturation have stronger effects on the platelet count and prothrombin time/INR.

**Table 7:**
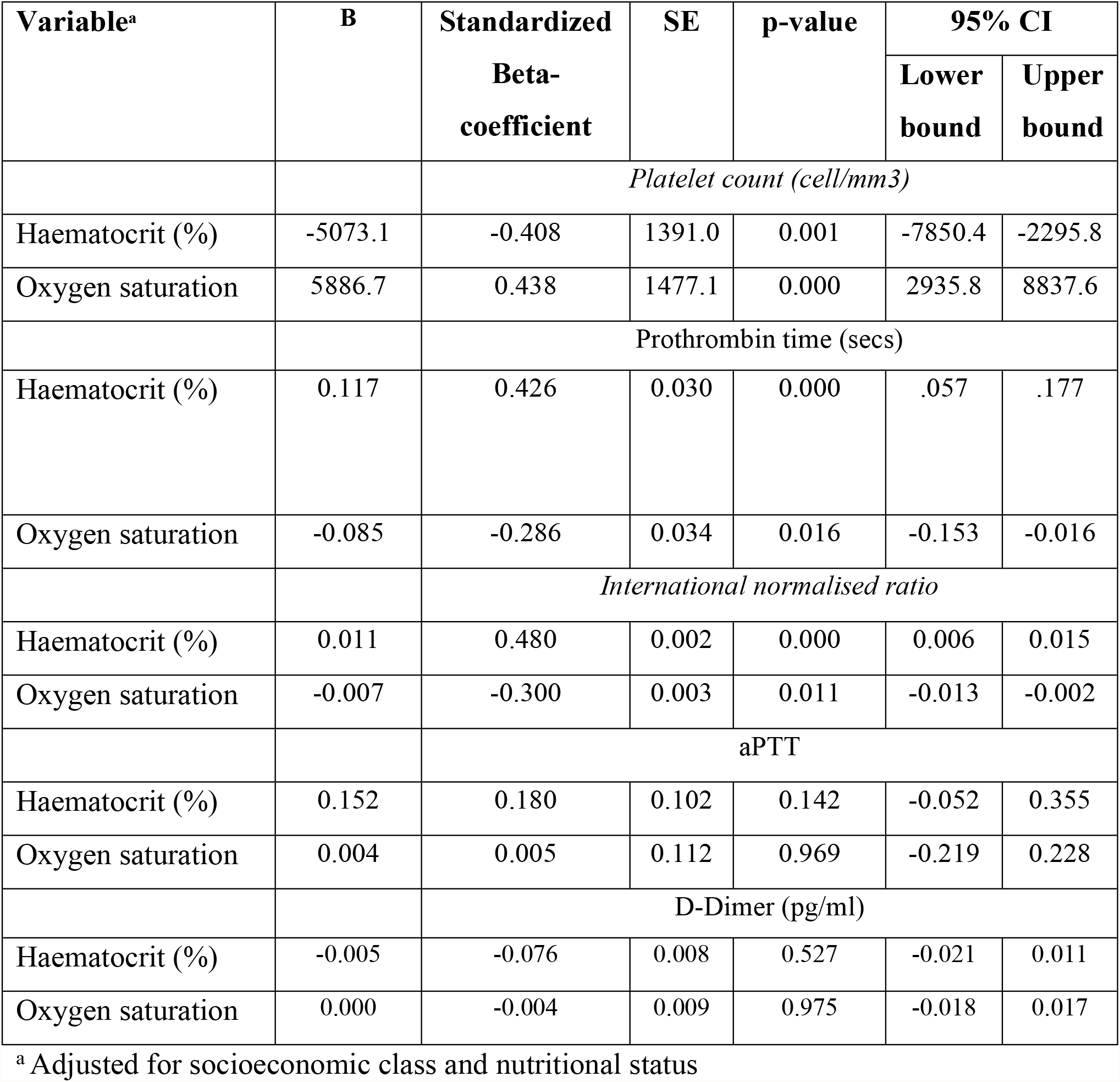
Multivariate logistic analysis of significant variables and Coagulation abnormality

Pearson’s correlation was carried out to demonstrate relationship between coagulation abnormality and factors such as age, anthropometry, oxygen saturation and haematocrit. Table 8 shows that haematocrit value and oxygen saturation had stronger effects on the platelet count and prothrombin time/INR. There was moderate negative correlation between haematocrit and platelet count (r = -0.414, p = 0.000) and a moderate positive correlation between the haematocrit level of children with CHD and prothrombin time/ INR (r = +0.416, p = 0.000). There was also a positive moderate correlation between the oxygen saturation level of children with CHD and platelet count (r = +0.443, p = 0.000). There was no correlation between age of patient, anthropometry and coagulation abnormality.

**Table 8:** Correlation between coagulation parameters and some independent factors

| **Variables (n=70)** | **Platelet**<br>**count** | **Prothrombin**<br>**time** | **Partial**<br>**thromboplastin**<br>**time** | **International**<br>**Normalised**<br>**ratio** | **D-dimer** |
| --- | --- | --- | --- | --- | --- |
| Age | $r = -0.215$<br>$p = 0.074$ | $r = 0.099$<br>$p = 0.415$ | $r = -0.068$<br>$p = 0.577$ | $r = 0.199$<br>$p = 0.099$ | $r = -0.324$<br>$p = 0.006$ |
| Weight for age | $r = -0.122$<br>$p = 0.340$ | $r = -0.143$<br>$p = 0.264$ | $r = -0.015$<br>$p = 0.908$ | $r = -0.146$<br>$p = 0.254$ | $r = -0.031$<br>$p = 0.811$ |
| Height for age | $r = -0.063$<br>$p = 0.604$ | $r = -0.076$<br>$p = 0.531$ | $r = -0.041$<br>$p = 0.735$ | $r = -0.087$<br>$p = 0.473$ | $r = -0.084$<br>$p = 0.489$ |
| BMI for age | $r = -0.104$<br>$p = 0.392$ | $r = -0.202$<br>$p = 0.093$ | $r = 0.044$<br>$p = 0.718$ | $r = -0.199$<br>$p = 0.098$ | $r = 0.000$<br>$p = 1.000$ |
| Oxygen saturation | $r = 0.443$<br>$p = 0.000$ | $r = -0.285$<br>$p = 0.017$ | $r = 0.000$<br>$p = 0.997$ | $r = -0.298$<br>$p = 0.012$ | $r = -0.024$<br>$p = 0.841$ |
| Haematocrit level | $r = -0.414$<br>$p = 0.000$ | $r = 0.416$<br>$p = 0.000$ | $r = 0.187$<br>$p = 0.122$ | $r = 0.471$<br>$p = 0.000$ | $r = -0.066$<br>$p = 0.586$ |

## Discussion

This study determined the prevalence of coagulation abnormalities (thrombocytopaenia, prolonged clotting profile and elevated d-dimer levels) in children with uncorrected congenital heart defects and reported a prevalence of 37.1%, compared to 7.2% in the healthy cohort. The mean clotting profile of children with uncorrected CHD was significantly prolonged and comparable to an earlier report by Cheung.[17] These findings in our study can be attributed to the uncorrected congenital heart defect, which has been reported to cause a chronic consumption coagulopathy.

Our prevalence was higher than that reported by Colon et al in the United States among a mixed cohort of adults and children, some of whom had corrected heart defects.[13] This is possibly due to the cohort of children with uncorrected CHDs in the current study, which reflects the pool of children with CHD in sub-Saharan Africa where cardiac intervention is delayed due to high cost of surgery and inadequate manpower. In comparison, our prevalence was lower than reports from the Indian sub-continent, which were in small patient populations. [12,18] These investigators proposed that their cohort had severe cardiac defects. Nevertheless, the relatively high prevalence of coagulation abnormality between CHD cohort in this current study compared to the healthy population further highlights that chronic disseminated intravascular coagulopathy occur in children with CHD. The prevalence among the apparently healthy population in this current study was comparable to the report by Alzahrani *et al* [19] in Saudi Arabia. It was however lower than figures reported by Acosta *et al* [20] in the US and Onakoya [21] in South West Nigeria respectively. The low rates in the current study might be linked to the exclusion of children with bleeding disorders as well as the study design utilized. It may then be assumed that coagulation disorders occur even among apparently healthy children. Viral infections have been implicated in the prevalence of coagulation defects in apparently healthy cohorts. This is due to the elaboration of cytokines that alter the balance between pro and anti-coagulation factors. Thus, pre surgical coagulation screening for all children should be advocated regardless of absence of a family history of bleeding disorders.

The prevalence of coagulation abnormalities in children with CCHD was significantly higher than the ACHD cohort, which is similar to earlier published reports. [13] In our population of children with CHD, slightly less than a quarter of the CCHD arm had thrombocytopenia with no thrombocytopenia reported among the ACHD arm. This is similar to a report in the US among a cohort of children with CCHD.[22] This was also corroborated by a study from Europe.[13] Decreased platelet counts may be due to the increased platelet destruction, aggregation and activation as well as reduced platelet production due to the right to left shunts, which deliver megakaryocytes into the systemic arterial circulation bypassing the lungs where megakaryocytic cytoplasm is fragmented into platelets.[23.24]

Similar to earlier reports, PT and aPTT were the most deranged coagulation screening tests in this study.[12,13] Prolonged PT/INR and APTT were significantly higher in children with CCHD compared to the ACHD cohort. A high prevalence (35.5% and 37.5%) of prolonged PT/INR in CCHD has also been reported among children with CCHD in East Africa. [25,26]. However, in a report from the United States, a higher rate was noted among children aged 3 to 19 months with CCHD. [27]. This was probably due to the predominance of infants in their study group, which may have partly accounted for the higher rates, as infants are known to have physiologic maturational delay in haemostasis.[28]

Elevated D-dimer level was seen equally in both ACHD and CCHD groups in the current study. A study in Nairobi however reported a significantly higher rate in the CCHD cohort.[26] Children with CCHD are at a higher risk of chronic thrombus formation due to hypoxic induced polycythaemia while, the chronic heart failure in ACHD causes cardiac dilatation, leading to reduced blood perfusion and possibly blood stasis with consequent tendency to thrombus formation.

Coagulation abnormalities were found to be highest in under fives, compared to the older age group. The difference was however not statistically significant. This finding is similar to the US study [27], who also found more coagulation abnormalities in participants less than 5 years. This suggests that this complication occur at an early age. Furthermore in the current study, there was no statistically significant relationship between coagulation abnormality and nutritional status.

Similar to published works, [12,13,17] our study showed a weak and negative correlation between oxygen saturation level and coagulation abnormalities. This relationship suggests that hypoxia contributes significantly to the initiation and sustenance of coagulation abnormalities as it leads to endothelial injury, and exposure of the sub-endothelial space to blood platelets, with subsequent activation of the coagulation cascade.[31] Hypoxia also results in hepatic hypo-perfusion and reduced production of clotting factors.

Furthermore, the correlation between haematocrit level, thrombocytopaenia and prolonged prothrombin time though weak, was statistically significant and in concordance with previous studies in Japan and United States of America. [22,32] This affirms the role of polycythaemia and platelet dysfunction in the promotion of abnormal coagulation cascade in uncorrected congenital heart defects. Elevated haemoglobin results in hyper-viscosity with resultant increased endothelial dysfunction, platelet activation and subsequently accelerated activation of the clotting cascade. The limitation in the present study was inability to use point of care testing for coagulation analysis, which is a more sensitive diagnostic test for coagulation analysis as it uses whole blood sample and better represent actual clotting function.

## Conclusion

It is evident that coagulation abnormalities are more prevalent among children with uncorrected cyanotic CHD when compared to those with acyanotic and apparently normal children. Oxygen saturation and haematocrit are factors associated with these abnormalities. There is thus a need for periodic evaluation for coagulation abnormalities in children with congenital heart defects to improve their quality of life and disease burden while they await corrective surgeries.

We recommend early and twice yearly regular screening for coagulation abnormalities as part of routine management of children with CHD.

## Data Availability

All relevant data are within the manuscript and its Supporting Information files.

## Acknowledgements

My sincere gratitude goes to all the children and the parents who participated in this study.

